# ctDNA detection by personalised assays in early-stage NSCLC

**DOI:** 10.1101/2021.06.01.21258171

**Authors:** Katrin Heider, Jonathan C. M. Wan, Davina Gale, Andrea Ruiz-Valdepenas, Florent Mouliere, James Morris, Nagmi R. Qureshi, Wendi Qian, Helena Knock, Jerome Wulff, Karen Howarth, Emma Green, Jenny Castedo, Viona Rundell, Wendy N. Cooper, Tim Eisen, Christopher G. Smith, Charles Massie, David Gilligan, Susan V. Harden, Doris M. Rassl, Robert C. Rintoul, Nitzan Rosenfeld

**Affiliations:** Cancer Research UK Cambridge Institute, University of Cambridge, Li Ka Shing Centre, Robinson Way, Cambridge CB2 0RE; Cancer Research UK Cambridge Centre – Cambridge, Cancer Research UK Cambridge Institute, Li Ka Shing Centre, Robinson Way, Cambridge CB2 0RE; Amsterdam UMC, Vrije Universiteit Amsterdam, Department of Pathology, Cancer Center Amsterdam, de Boelelaan 1117, Amsterdam, The Netherlands; Royal Papworth Hospital NHS Foundation Trust, Cambridge CB2 0AY, UK; Cambridge Clinical Trials Unit, University of Cambridge School of Clinical Medicine; Cambridge Clinical Trials Unit – Cancer Theme, Cambridge, UK; Inivata Ltd, Glenn Berge Building, Babraham Research Campus, Cambridge CB22 3FH, UK; Addenbrooke’s Hospital, Cambridge CB2 0QQ, UK; Department of Oncology, University of Cambridge Hutchison–MRC Research Centre, Box 197, Cambridge, Biomedical Campus, Cambridge CB2 0XZ, UK

## Abstract

Blood-based assays have shown increasing ability to detect circulating tumour DNA (ctDNA) in patients with early-stage cancer. However, detection of ctDNA in patients with non-small cell lung cancer (NSCLC) has continued to prove challenging. We performed retrospective analysis to quantify ctDNA levels in a cohort of 100 patients with early-stage NSCLC prior to treatment with curative intent enrolled in the LUCID study (NCT04153526). Where tumour tissue was available for whole exome sequencing, mutations identified were used to define patient-specific sequencing assays. For those 90 patients, plasma cell-free DNA was sequenced to high depth across capture panels targeting a median of 328 mutations specific to each patient. Data was analysed using Integration of Variant Reads (INVAR), detecting ctDNA in 66.7% of patients, including 52.7% (29 of 55) patients with stage I disease and >88% detection for patients with stage II and III disease (16/18 and 15/17). ctDNA was detected in plasma at fractional concentrations as low as 9.1×10^−6^, and in patients with tumour volumes as low as 0.23 cm^3^. A 36-gene sequencing panel (InVisionFirst-Lung™) was used to analyse plasma DNA in 27 samples including the 10 cases without tumour exome data, and detected ctDNA in 59% of samples tested (16 of 27). Across the entire cohort, detection rates were higher in squamous cell carcinoma patients compared to adenocarcinoma patients (81% vs. 59%). Detection of ctDNA prior to treatment was associated with significantly shorter time free from relapse, across all patients and in patient subgroups, with Hazard Ratios >11 for selected patient subsets. Our analysis indicates that for patients with stage I NSCLC, the median ctDNA fraction in plasma is approx. 12 parts per million (0.0012%). This indicates the limits of detection that would be required for ctDNA-based liquid biopsies to detect ctDNA in the majority of patients with early-stage NSCLC.

## Introduction

Early detection of cancer using circulating tumour DNA (ctDNA) is an area of intense current investigation. Non-invasive methods to diagnose cancer earlier will provide patients more treatment options and a better chance of survival. ctDNA analysis can identify DNA from cancer cells in the plasma, and can provide a unique opportunity to diagnose patients in a minimally invasive way *(1)*. Non-small cell lung cancer (NSCLC) is one of the leading causes of cancer death, in part due to the large number of patients presenting with late-stage disease. The SUMMIT trial, launched in April 2019, aims to clinically evaluate an assay for detection of cancer through analysis of circulating cell-free DNA (cfDNA) in plasma, in a population at risk for lung cancer *(2)*. However, in patients with early-stage disease, ctDNA levels are low, and detection of early-stage NSCLC with current methods has had limited sensitivity.

In the recent published literature, reported detection rates of ctDNA in patients with early-stage NSCLC have ranged between 15% and 45% *(3–7)*. Low levels of ctDNA in early stage NSCLC, which often fall below the limits of detection of most platforms used to date, may partly be due to low disease volume *(7, 8)*. Additionally, it may also be impacted by differences in tumour biology whereby more rapidly proliferating, aggressive or advanced-stage cancers may release ctDNA at higher rates, and in sufficient amounts to be detected more readily in plasma *(5, 9)*. Previous studies have used patient-specific analysis of ∼20 mutations per patient *(7)*, fixed panels ranging in size from 2kb to 188kb and covering 16-128 genes *(3, 4, 6)*, or a combination of fixed and patient-specific panels *(5)*. Some assays leverage patterns of ctDNA coverage or fragmentation *(10, 11)*, epigenetic changes such as methylation *(12–14)*, or combine information from additional analytes and tumour markers *(4, 15)*.

In this study we aimed to quantify ctDNA fractions in plasma of 100 patients with early-stage, treatment-naïve NSCLC enrolled in the LUng Cancer CIrculating Tumour Dna Study (LUCID, NCT04153526) to define the distribution of ctDNA levels in this setting. Where tumour tissue was available (n=90), we designed patient-specific sequencing panels and employed our previously published Integration of Variant Reads (INVAR) analysis pipeline *(16)*. INVAR utilises large, patient-specific mutation lists in combination with noise reducing methods, signal-weighting and signal integration to detect and quantify low ctDNA fractions. We also applied a commercial targeted sequencing assay *(17, 18)* to detect and quantify ctDNA in samples collected before treatment from 27 patients who underwent radiotherapy ± chemotherapy, including those patients for whom tumour tissue was unavailable (n = 10, Fig. 1A). Data from both methods was used to assess ctDNA detection rates and ctDNA fractions, and their differences between cancer stages and histological subtypes.

**Fig. 1:**
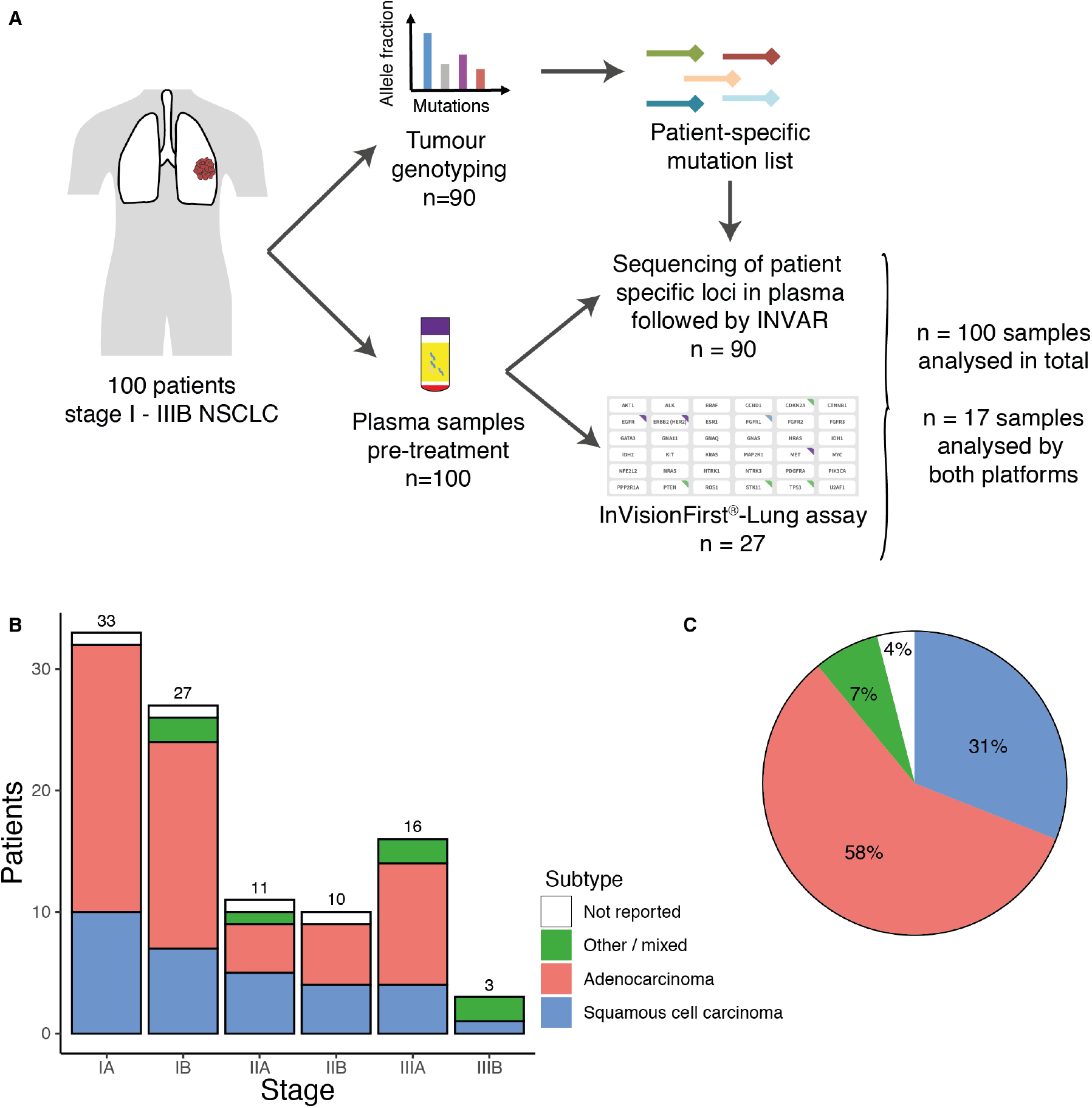
Study design and distribution of tumour stages and subtypes. (A) 100 patients eligible for treatment with curative intent by either surgery (n=70) or chemo-radiotherapy (C-RT, n=30) were recruited to the LUCID study. From each patient, a plasma sample was collected prior to initiation of treatment. Furthermore, a tumour specimen was collected from all of the surgical patients and from 20 of the C-RT patients. Where tumour tissue was available (n=90), it was analysed by whole exome sequencing and identified mutations were used to design patient-specific hybrid capture panels for targeted sequencing. Custom capture sequencing data was generated and analysed using the INVAR pipeline (n=90) *(16)*. For 27 of the C-RT patients, including all 10 patients for whom tumour tissue was unavailable, plasma samples were analysed using the InVisionFirst^®^-Lung assay. 17 of the samples were analysed by both platforms. (B) Distribution of disease stage at diagnosis for 100 patients in the LUCID cohort. 60% of the patients were diagnosed with stage I disease at presentation. (C) Subtype distribution in the LUCID cohort.

## Results

A total of 100 treatment-naïve patients with stage I–IIIB NSCLC undergoing surgery or radiotherapy ± chemotherapy were recruited to the LUCID study (“Lung Cancer circulating tumour DNA study”). The cohort predominantly consisted of patients with stage I disease at diagnosis (n=60), with 21 and 19 patients with stage II and III disease stage at diagnosis respectively (Fig. 1B). The median age of patients in the cohort was 72 years (range: 44 – 88 years), and 89 (90%) were current or previous smokers. Tumour subtype information was available for 96 patients and consisted of 32% (31/96) squamous cell carcinoma, 60% (58/96) adenocarcinoma and 7% (7/96) other subtypes (Fig. 1C). Patients either underwent surgery (70/100) or radiotherapy ± chemotherapy (30/100). Plasma samples obtained before the initiation of treatment were analysed using one or two methods. For 90 patients, we obtained a tissue sample, either from surgery or remaining material from the diagnostic biopsy, and used tumour whole exome sequencing information to guide ctDNA analysis in plasma using patient-specific sequencing panels and INVAR (**Methods**). Additionally, we applied the InVisionFirst^®^-Lung assay to 27 plasma samples, including 10 cases where no tumour tissue was available and 17 samples with available tumour tissue that were also analysed using INVAR.

### Mutational landscape in NSCLC

For 90 patients with available tissue, we performed whole exome sequencing of matched tumour and buffy coat material. Using a combination of mutation callers (**Methods**) we identified a median of 328 mutations per patient (IQR 205 to 491, total 44,514 mutations). We leveraged these mutations to analyse commonly mutated lung cancer genes and mutation signatures in this cohort.

First, we assessed the frequency of mutations in our cohort in genes that were previously identified as being frequently mutated in both adenocarcinoma and squamous cell carcinoma lung cancers patients *(19, 20)*. For patients with adenocarcinoma, the genes most commonly containing mutations were KRAS (43.6%) and TP53 (36%, Fig. 2A). In the literature KRAS and TP53 mutations were observed in 33% and 46% of patients, respectively *(19)*. TP53 was the most commonly mutated gene in the squamous cell carcinoma cohort, with over 64% of patients having an alteration (Fig. 2B), while the literature describes TP53 mutations in 81% of patients *(20)*.

**Fig. 2:**
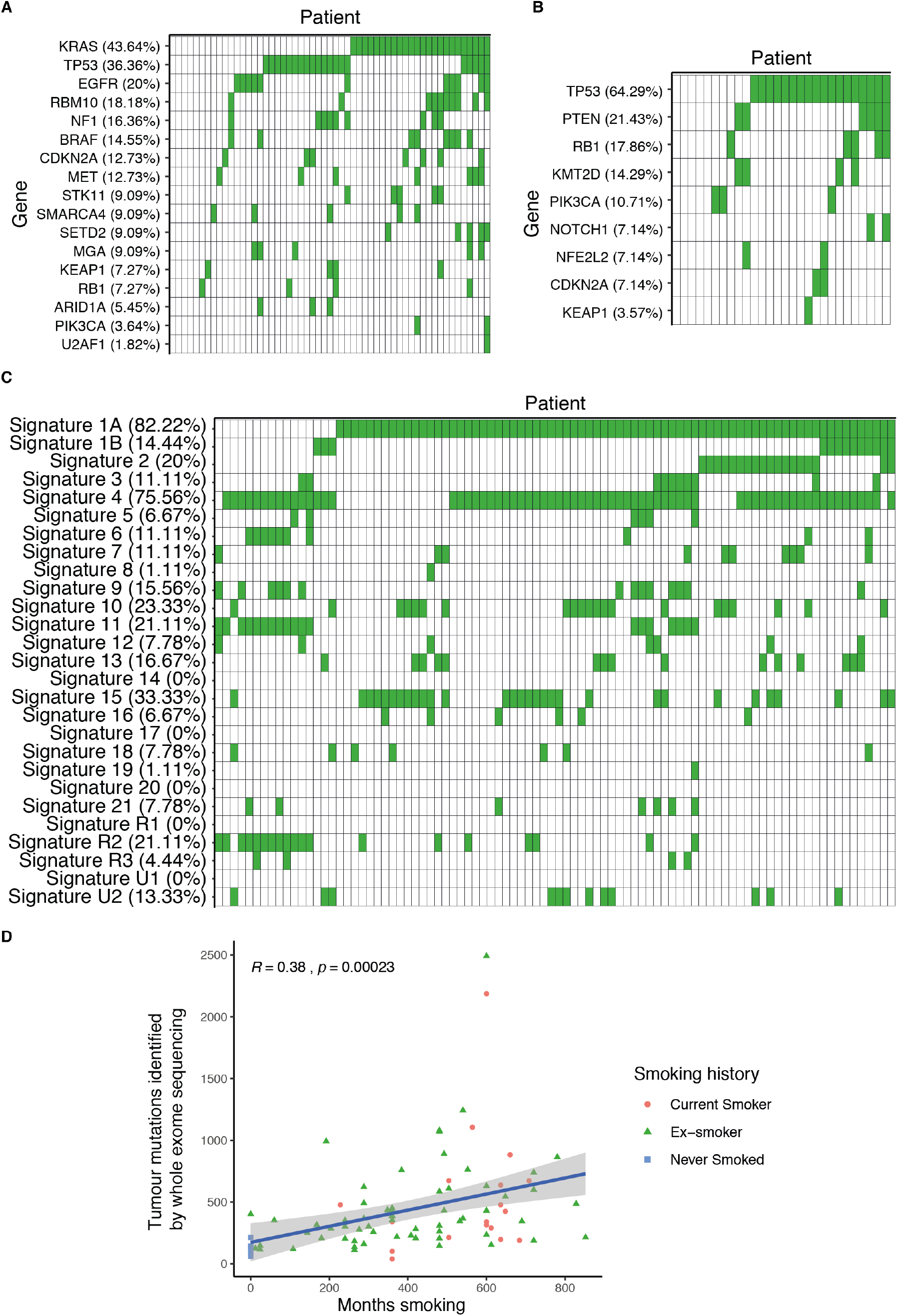
Analysis of tumour whole exome sequencing data in NSCLC. (A) Genes commonly mutated in adenocarcinoma were identified as being significantly mutated from Cancer Genome Atlas (CGA) analysis *(19)*. Mutations in these genes were identified in the present cohort (filled boxes) from whole exome sequencing data, for patients with adenocarcinoma and where tumour tissue WES was available (n=55 patients). (B) Genes commonly mutated in squamous cell carcinoma were identified as significantly mutated from CGA analysis *(20)*. Mutations in these genes were identified in the present cohort (filled boxes) from whole exome sequencing data, for patients with squamous cell carcinoma and where tumour tissue WES was available (n=28 patients). (C) Mutation signatures were obtained using deconstructSigs *(22)* and their presence is shown (filled boxes) for the 90 patients in this study from whole exome sequencing data, for the patients where tissue WES data was available. (D) A significant correlation was observed between the tumour mutation burden and the extent of smoking history (Pearson’s R = 0.38, p = 0.00023).

We further analysed mutation signatures, assessing the distribution of mutations in different trinucleotide contexts *(21)*. Using the deconstructSigs package *(22)*, we applied the 26 signatures of mutation processes identified by Alexandrov and colleagues *(21)* to our cohort (Fig. 2C). The two most commonly observed signatures were signature 1A and signature 4. The ageing-related signature 1A *(21)* was most abundant (82%) and fits with the median patient age of 72 in this cohort. A majority of patients also showed the smoking-related signature 4 (76%) (Fig. 2C) *(21)*, in agreement with the high rate (90%) of current and previous smokers in the cohort.

Finally, we compared the tumour mutation burden with smoking history. Both current and previous smokers had a significantly higher tumour mutation burden (TMB) compared to never smokers, confirming previous observations (fig. S1A) *(23)*. We observed a significant correlation when comparing the TMB with the number of months smoked for each patient (Pearson’s R = 0.38, p = 0.00023, Fig. 2D)*(23)*. In contrast, we did not observe a significant correlation of TMB with age (fig. S1B).

### Detection of ctDNA using patient-specific sequencing panels and the INVAR pipeline

Patient-specific mutation lists, generated through tumour and buffy coat whole exome sequencing, were used to design three panels of oligonucleotide baits for hybrid-capture, that covered 9,831, 16,227 and 18,456 mutation loci each, identified in 19, 34 and 37 patients respectively. These targeted a median of 328 mutant loci per patient across the cohort of 90 patients. Cell-free DNA was extracted from plasma samples collected prior to treatment for each of the 90 patients, and a sequencing library was created from a mean of 13.6 ng (IQR: 14.7 ng - 15 ng) of cfDNA from each sample, equivalent to approximately 4500 copies of the genome. Genomic regions of interest were selected from each library by hybrid capture using the appropriate panel of oligonucleotides that included the mutant loci identified in tumour analysis for that patient (**Methods**). Libraries were sequenced to a median of 67,696,747 total reads per library (IQR 59,262,532 – 73,914,836) across patient specific panels with sizes of 2.14, 2.65 and 2.99 Mbp, respectively.

Sequencing data were analysed using the INtegration of VAriant Reads (INVAR) analysis pipeline, as previously described *(16)*. Briefly, read families were generated based on unique molecular identifiers to reduce sequencing errors; probabilities were assigned to each variant read based on error rates that were calculated according to trinucleotide context; probability weights were assigned based on the mutation allelic fraction in the original tumour sample and the fragment length of the present cfDNA molecules; and an INVAR score was calculated for each sample by integrating signal across the panel. Resulting INVAR scores were compared between patients and controls to determine presence of ctDNA. In this cohort, 99.8% of mutations were private to one patient. This minimal overlap allows us to use data from individual patients as a control for other patients that were captured using the same custom panel. This maximised the use of sequencing data and increased the number of control samples without having to sequence additional samples *(16)*. For each sample, an integrated mutant allele fraction (IMAF) was calculated to estimate the fraction of DNA originating from the tumour, as the depth-weighted average of the mutation rates at each of the loci corrected for the background error rate at each locus.

For each of the three sequencing panels, an INVAR score threshold was determined for detection of ctDNA using receiver operating characteristic (ROC) analysis to maximise the sensitivity and specificity (averaged with equal weights). For each panel, we required a specificity of at least 95% (Fig. 3A). Applying the same detection thresholds to sets of samples from individuals without cancer that were captured using the same panels, we observed a mean overall specificity of 95.63% (Fig. 3A) with observed specificities of 97.4%, 95.5% and 94% for the individual panels. Using those INVAR score specificity thresholds, ctDNA was detected in 66.7% (60 of 90) of samples. Amongst the patients with detected ctDNA a median of 2.6% (IQR 0.7% - 24.7%) of the mutant loci targeted for sequencing showed somatic signal. In these patients a median of 9 loci had signal in plasma (IQR 2 – 43). In comparison, a fixed gene panel designed for detection of ctDNA in NSCLC, covering 125 kb and 139 genes commonly mutated in NSCLC *(24)*, would cover only 336 (0.75%) of the 44,513 mutations analysed in plasma in this study using tumour-informed assays.

**Fig. 3:**
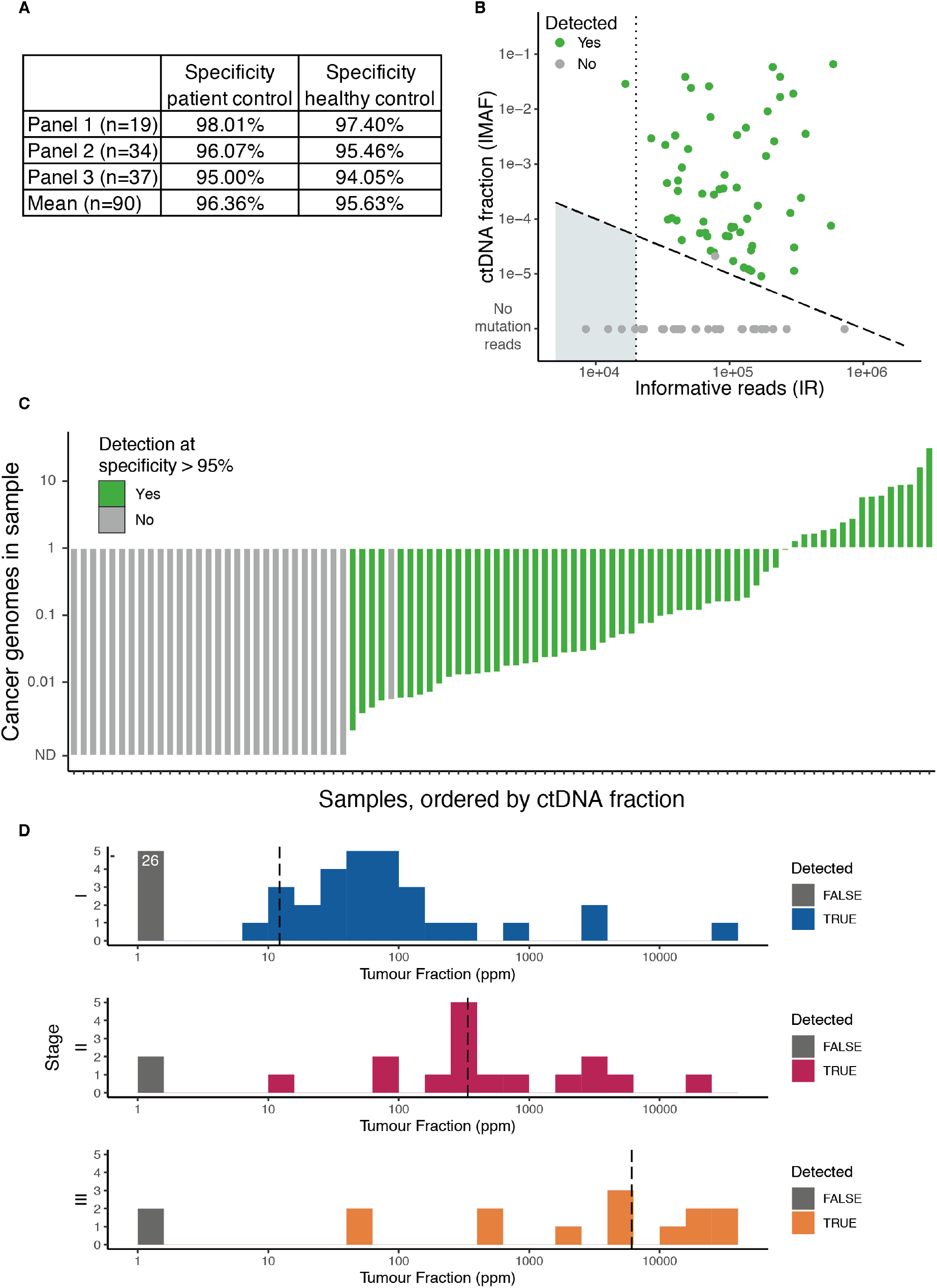
ctDNA detection in early-stage NSCLC by patient-specific sequencing panels and INVAR analysis. (A) Specificities for the three custom capture sequencing panels in this study. Specificities were assessed twice. Once using other (non-matched) patients as controls, and once using an independent set of healthy controls. (B) Informative Reads (IR) are plotted against the Integrated Mutant Allele Fraction (IMAF) which estimates the ctDNA levels in each sample. Sensitivity of the INVAR pipeline increases with increasing IR and the threshold for ctDNA detection can be roughly estimated as 1/IR (indicated by dashed diagonal line). Samples with less than 20,000 IR (dark grey box) allow only for limited detection sensitivity. Samples in which ctDNA was detected by INVAR are shown in green. Undetected samples are shown in grey. (C) Cancer genomes detected in the analysis of plasma samples. The total number of cancer genomes represented in each sample analysed was estimated for patients with detected ctDNA. In many samples, ctDNA was detected even though less than 0.1 of a single cancer genome was present, and would likely not be detected without the analysis of a greater number of confirmed markers. (D) ctDNA tumour fractions, in parts per million (ppm), detected in baseline plasma samples according to disease stage at diagnosis, for the 90 patients analysed with INVAR (55, 18 and 17 patients with stage I, II and III disease respectively). Median ctDNA levels per stage are indicated with vertical dotted lines.

We previously showed that we could estimate the limit of sensitivity in each sample based on the number of informative reads (IR), which are individual sequence reads that overlap with patient-specific mutation loci *(16)*. The number of informative reads further provides an estimate of the sensitivity limit in each sample (roughly equivalent to 1/IR) *(16)*. We generated a median of 87,523 Informative Reads (IR) for each sample at patient-specific mutated loci (IQR 44,149 – 156,436). For 5 of the 90 samples, less than 20,000 IR were obtained, limiting our ability to detect ctDNA at fractions below 5×10^−5^ (Fig. 3B, first dotted line). Despite this limited sensitivity, for one patient with stage IIIA disease, we detected ctDNA was detected at an allelic fraction of 0.029. In the four remaining samples ctDNA was not detected with the available sequencing data, but may potentially be detected if additional sequencing was performed to generate more IR, thus increasing sensitivity. Additionally, the INVAR pipeline infers the total cancer (haploid) genomes in the sequencing data analysed by dividing the total number of unique mutated molecules by the total number of unique cfDNA molecules sequenced across the mutated loci *(16)*. For the 60 samples with ctDNA detected using INVAR we inferred a median number of (haploid) cancer genomes of 0.07 (IQR 0.02 to 1.02) (Fig. 3C), meaning that the sequencing data for most samples represented less than 10% of a cancer cell’s (haploid) genome. Across the 90 patients, we established the distribution of pre-treatment ctDNA levels in patients presenting with NSCLC of different stages (Fig. 3D). ctDNA was detected in >50% of patients in each of the stages: 29/55 (52.7%) for stage I, 16/18 (88.9%) for stage II, and 15/17 for stage III (88.2%). This allowed the evaluation of an estimated median fraction of ctDNA in each stage: 12 parts per million (ppm), 338 ppm and 7,419 ppm (0.0012%, 0.034% and 0.74%) in stages I, II and III respectively (Fig. 3D).

### Detection of ctDNA using InVisionFirst^®^-Lung assay

Next, we applied the InVisionFirst^®^-Lung assay (Inivata Ltd.) *(17, 18)* to analyse 27 of the 30 pre-treatment plasma samples from patients undergoing radiotherapy ± chemotherapy. Ten of these had no tumour tissue available, and thus patient-specific hybrid capture panels could not be designed. The remaining 17 samples, selected based on availability of plasma and blinded to other data, had tumour tissue available and were analysed by both methods. The InVisionFirst^®^-Lung assay *(17)* covers 10.61kb and detects single nucleotide variants, copy number variants and insertions and deletions in regions from 36 selected cancer-related genes. Based on the TAm-Seq method originally described by Forshew et al *(25)*, this enhanced TAm-Seq (eTAm-Seq™) technology amplifies DNA fragments of length 72bp-154bp in a two-step multiplex PCR amplification to prepare amplicon sequencing libraries *(17)*. In a validation study the assay demonstrated 99.48% sensitivity and 99.99% specificity for mutations with allelic fractions ranging from 0.25% to 0.33%, the recorded limit of detection. For mutations with allelic fractions between 0.13% and 0.16% as well as 0.06% and 0.08%, the assay reached detection rates of 88.93% and 56.25%, respectively *(18)*.

Using a median input of 3.7mL of plasma (3.2mL – 4mL) and a median of 7,240 copies of the genome (2,720 – 16,000), this non-patient-specific approach that does not require matched tumour or normal DNA initially detected ctDNA in 17 of 27 patients. For 12 of these 17 patients, tumour and buffy coat data was available and its analysis led to reclassification of one mutation as a germline mutation (signal observed in the matched buffy coat). No tumour tissue was available for the other five patients with detected mutations. Overall, after the exclusion of the aforementioned germline variant, the InVisionFirst^®^-Lung assay detected ctDNA in 16 of 27 patients (59%) with a mean of 1.67 alterations per patient and a total of 27 mutations across the cohort. 18 of these mutations (66.7%) were detected at an allelic fraction lower than 0.01, and 12 (44.4%) were detected at an allelic fraction below 0.005. *TP53* was the most commonly mutated gene in these patients (37%), followed by alterations in *KRAS* (22%), *STK11* (11%), *CDKN2A* (11%), *PTEN* (4%), *NFE2L2* (4%), *KIT* (4%) and *PIK3CA* (4%) (Fig. 4A). The InVisionFirst^®^-Lung assay detected ctDNA in 73% of the samples from patients with stage III disease (11 out of 15). Detection rates decreased to 50% (3 out of 6) in stage II patients and 33% (2 out of 6) in stage I patients.

**Fig. 4:**
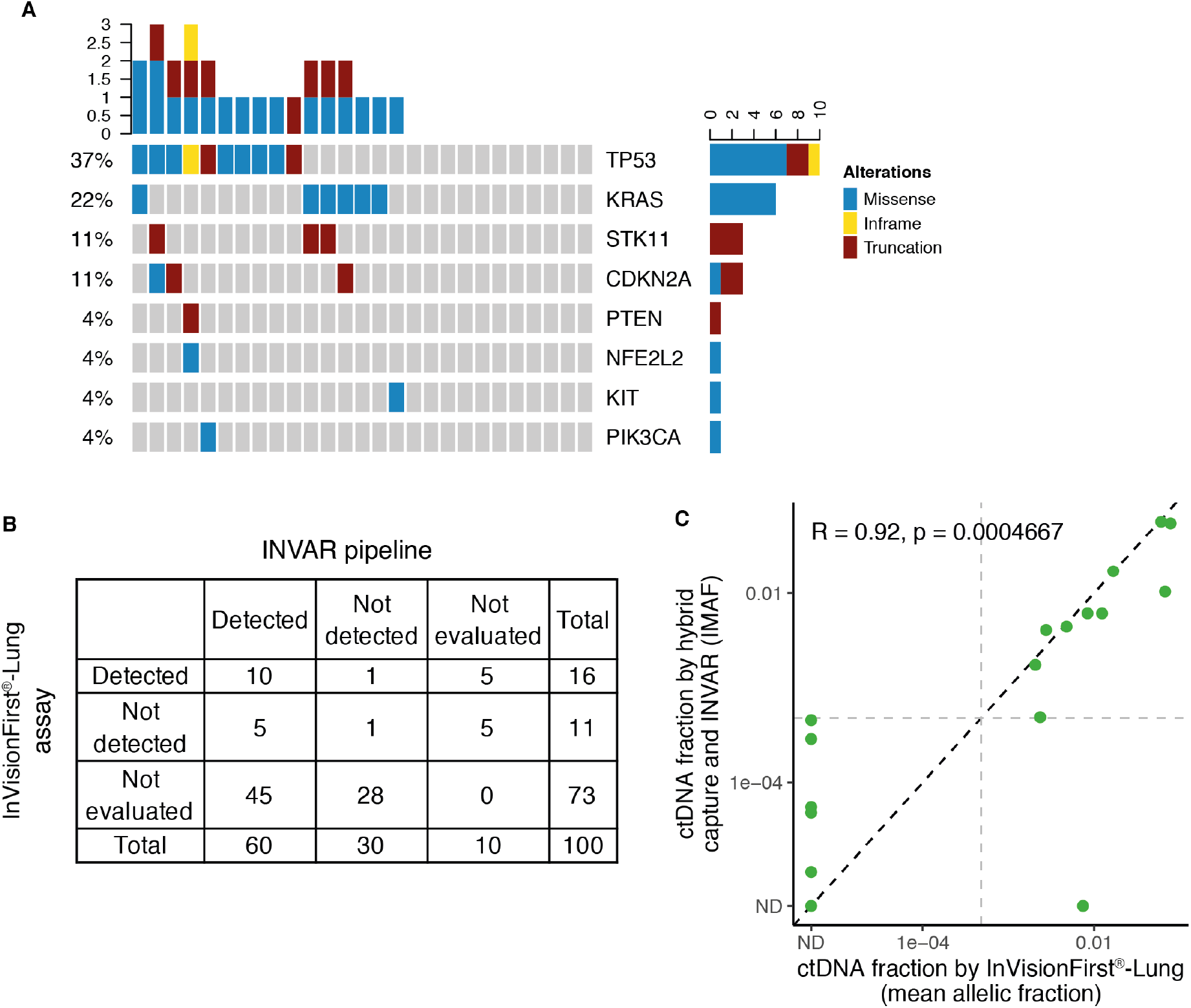
InVisionFirst^®^-Lung assay analysis in 27 patients with NSCLC. (A) A total of 27 mutations were identified; most alterations were identified in *TP53* (37%), followed by *KRAS* (22%). Where matched tissue data was available, we confirmed 70% (14/20) of these mutations. Of the remaining 6 mutations, one was confirmed to be a germline variant, two splice variants were not covered in our analysis, and three were not detected in tumour whole exome sequencing. (B) Comparison of ctDNA detection between INVAR and InVisionFirst^®^-Lung. Sensitivity of INVAR alone was 66.7% (60/90) while the InVisionFirst^®^-Lung assay detected ctDNA in 59% (16/27) of samples. 17 samples were analysed by both platforms with a concordance in ctDNA detection of 64.7% (11/17, 10 samples detected and one sample undetected by both platforms). (C) ctDNA fraction correlation between INVAR and InVisionFirst^®^-Lung. 10 samples were detected by both platforms; yielding a correlation in ctDNA fraction of 0.92 (Spearman’s r, p = 4.7×10^−4^).

Plasma samples for 17 patients were analysed with both the InVisionFirst^®^-Lung assay and the INVAR pipeline. Amongst these samples, 10 (58.8%) were detected by both methods and one (5.9%) was detected by neither method (Fig. 4B). When comparing the ctDNA fractions of the 10 cases detected by both methods (using the mean ctDNA fraction for samples with more than one mutation detected with the InVisionFirst^®^-Lung assay), we observed a correlation of 0.92 (Spearman’s r, p = 4.7×10^−4^, Fig. 4C). Of the remaining discordant samples (n=6), ctDNA was detected in 5 samples using patient-specific INVAR analysis, at median IMAF of 5.5 ×10^−5^, but not detected by with the InVisionFirst^®^-Lung assay (Fig. 4B). There was one case in which ctDNA was detected with a mean ctDNA fraction of 0.007 with the InVisionFirst^®^-Lung assay but not detected by the INVAR pipeline (Fig. 4C). This case had only 40 patient-specific mutations identified by whole exome sequencing and capture sequencing generated12,391 IR, thereby limiting the potential sensitivity of INVAR, which performs better with larger mutation lists and more IR *(16)*.

### Overall detection rates in NSCLC

In this cohort of patients with NSCLC treated with curative intent, ctDNA was detected by one or both methods in plasma samples collected prior to treatment in 66 of the 100 patients (66%). We evaluated ctDNA detection by stage and further annotated the samples with demographic variables (Fig. 5A). We observed detection rates of 51.7% for ctDNA in patients with stage I disease, 85.7% for patients with stage II disease, and 89.5% for patients with stage III disease (Fig. 5A, fig. S2A). Similar to previous reports on ctDNA detection in NSCLC, we noted the highest detection rates and ctDNA levels for the advanced stage patients and observed similar or higher detection rates compared to other reports (Fig. 5B, fig. S2A) *(3–7)*. We quantified ctDNA down to a mutant allele fraction of 9.1×10^−6^. Overall, the median detected allelic fraction across all patients was 3.2×10^−4^ (IQR 5.6×10^−5^ to 3.4×10^−3^, Fig. 5A).

**Fig. 5:**
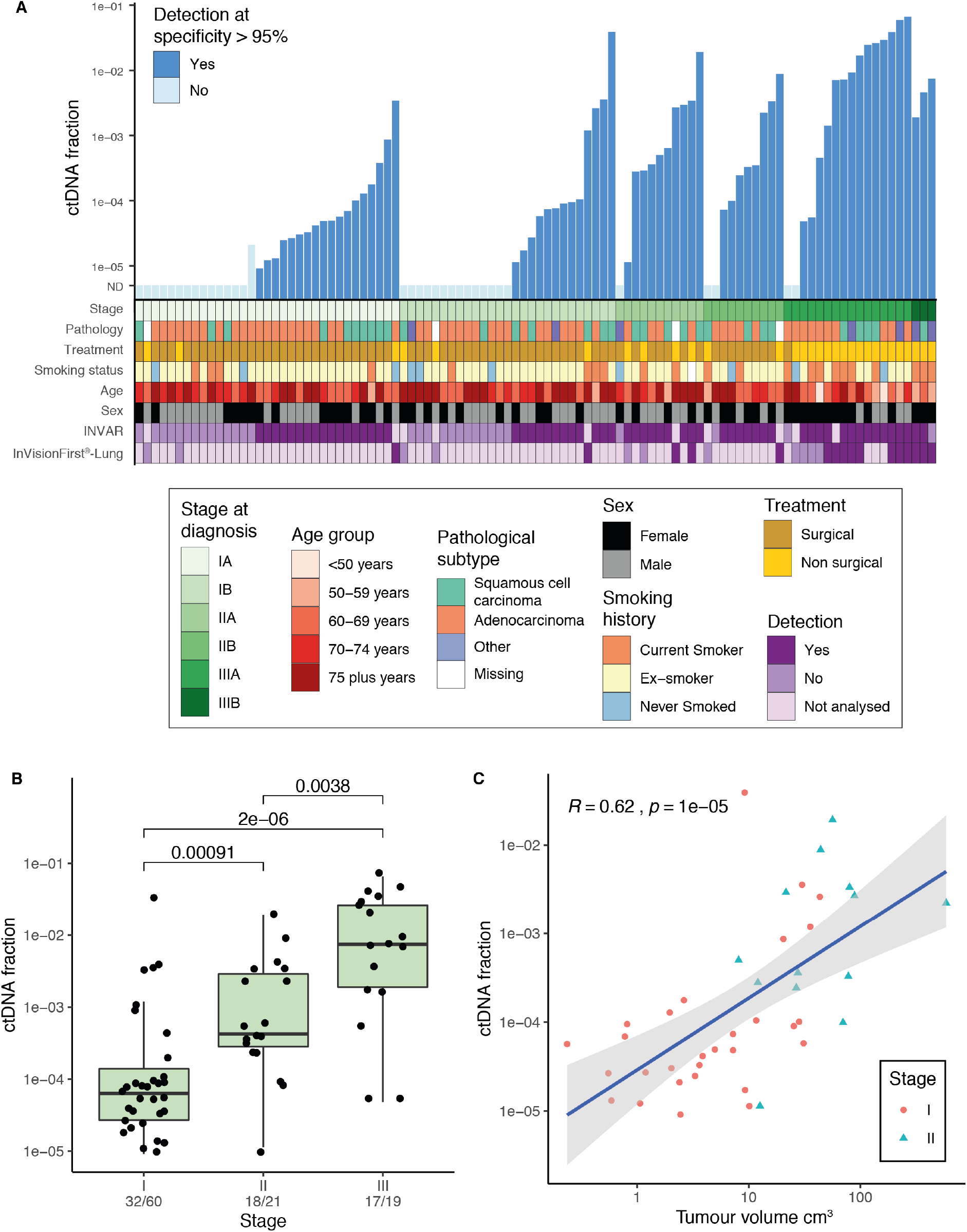
Summary of ctDNA levels and patient demographics. (A) In total, ctDNA was detected in 66 of 100 samples by at least one of the two analysis approaches. Patients are ordered by stage at diagnosis and detection/ctDNA levels; patient demographics are indicated below their respective ctDNA fraction. Detection rates are reported in the text. (B) Boxplot of ctDNA levels by stage, showing only cases in which ctDNA was detected. Number detected/number tested are indicated on the figure for each stage. (C) Correlation between ctDNA and tumour volume for patients with stage I disease (red dots) and stage II disease (turquoise triangles) in cases where ctDNA was detected and tumour measurements were available (n=44). Pearson’s R = 0.62, p = 1×10^−5^.

Histological subtype information was available for 96 of the 100 patients. Similar to previous reports *(5, 7)*, we observed higher detection rates of ctDNA in patients with squamous cell carcinoma compared to adenocarcinoma, with detection in 81% (25 out of 31), and 59% (34 out of 58) of patients, respectively. ctDNA was detected in 71% of patients with other subtypes (5 out of 7). Comparing ctDNA levels in detected samples, we saw significantly lower levels in patients with adenocarcinoma compared to other subtypes (fig. S2B).

Using our patient data, we explored the change in ctDNA detection by stage when applying varying detection thresholds. While detection rates for both stage II and III patients only started to drop off slightly at a detection threshold of 1×10^−4^ (almost 100-fold higher than the lowest tumour fraction observed in this study), detection rates in stage I patients decreased by more than half at the same threshold (fig. S2C), highlighting the need for a very sensitive assay for the detection of ctDNA in patients with early stage cancers.

We obtained tumour volumetric data for 41 of the 49 patients with stage I and II disease in which ctDNA was detected prior to treatment. The correlation between ctDNA concentration and tumour volume (cm^3^) was significant with a correlation of 0.62 (Pearson’s r, p = 1×10^−5^, Fig. 5C). We compared our correlation between ctDNA and tumour volume with previous studies in the field of lung cancer and found a similar significant strong correlation between ctDNA and tumour volume in this study, with lower ctDNA fraction for the same tumour volume observed in this study (fig. S3A) *(5, 7, 26)*. Additionally, INVAR was able to detect ctDNA down to a tumour volume of 0.23 cm^3^, a lower volume compared to previous studies (fig. S3A). Analysing the relation between tumour volume and ctDNA fractions separately for different histological subtypes (fig. S3B), we found that patients with adenocarcinomas had lower ctDNA fractions compared to patients with squamous cancers, for a given disease volume.

### ctDNA detection pre-treatment and patient outcomes

We analysed patient survival and relapse outcomes using using the Kaplan-Meier approaches, compared them between different groups using the Log-rank tests, and estimated hazard ratio using the proportional hazard (Cox) models (Fig. 6 and fig. S4). We analysed outcomes across all patients, and in patient subgroups, using several outcome metrics. Overall survival (**OS**) counted any death as an event, and patient data were right-censored if they were lost to follow-up (all remaining patients were right-censored at the time of the study close). As ctDNA analysis by INVAR was based on detection of mutations specific to the patient’s primary tumour, we evaluated freedom from relapse (**FFR**) as the time that patients remained free from relapse of the original primary tumour. In this analysis we considered as events only clinical relapse of the original primary tumour, and patient data was right-censored when patients presented with a second primary cancer that progressed, upon death from causes other than relapse of the first primary disease, or at loss of follow up/study close. We further analysed cancer-free survival (**CFS**), which evaluated the time until any cancer or death events, whichever occurred first, and patient data were right-censored at loss of follow-up or study close.

**Fig. 6:**
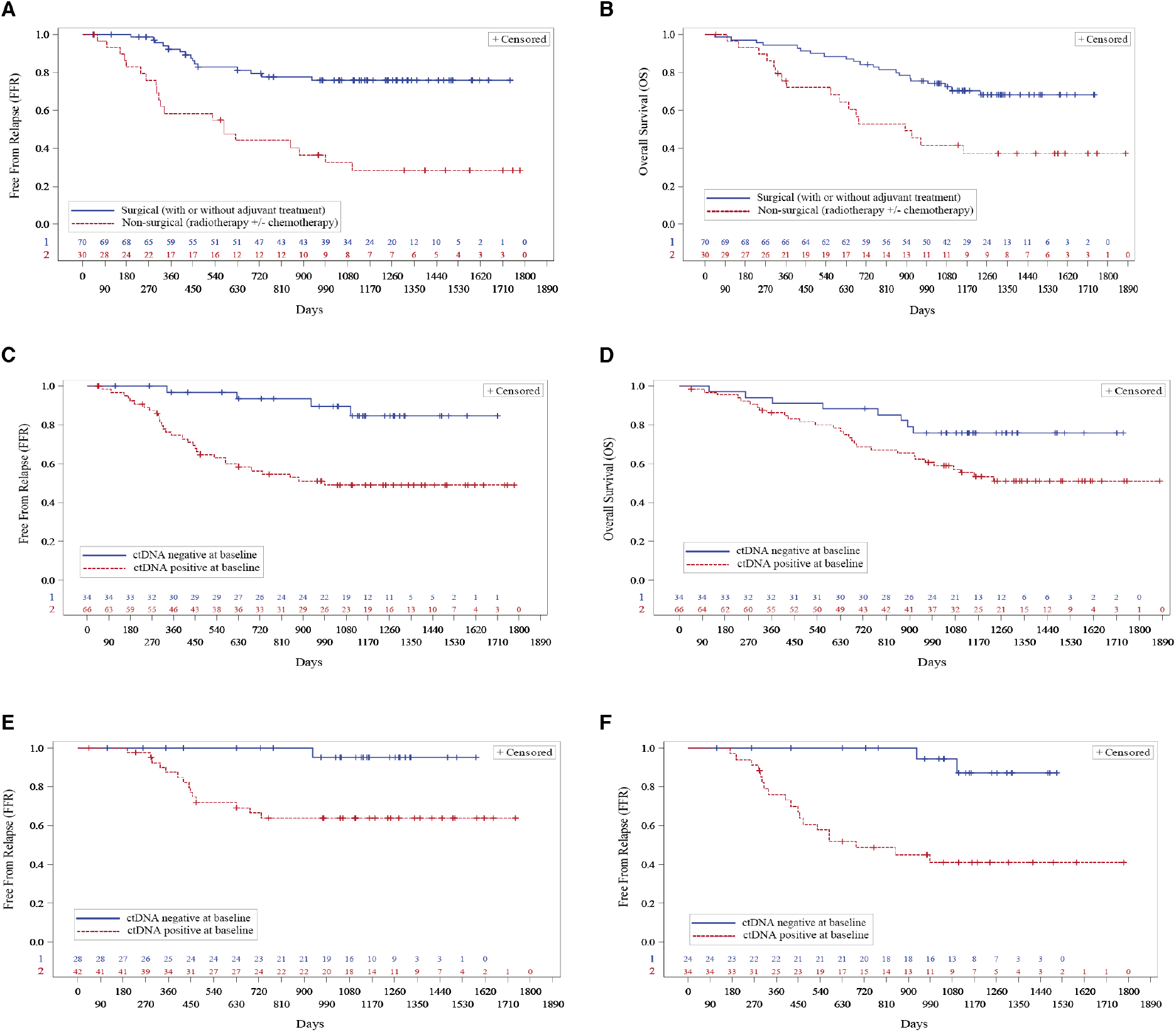
Analysis of patient outcomes and ctDNA detection. Kaplan-Meier plots showing overall survival (**OS**) and the time that patients remained free from relapse of the original primary tumour (**FFR**). FFR analysis considers as events only clinical relapse of the original primary tumour, and patient data is right-censored when patients present with a second primary cancer that progresses or upon death from causes other than relapse of the first primary disease. Analyses of cancer-free survival considering all cancer or death events (**CFS**), and additional OS analysis, are presented in fig. S4A and B) Comparison of FFR time (A) and OS (B) for patients who underwent non-surgical treatment (red, dotted) vs. surgical treatment (blue). Median FFR time of 578 days vs. not reached (Log-Rank Test p-value <0.0001); HR for non-surgical treatment: 4.35, p-value <0.0001). Median OS time of 893 days vs. not reached (Log-Rank Test p-value 0.002); HR for non-surgical treatment: 2.71, p-value 0.002). C+D) Comparison of FFR time (C) and OS (D) for patients with ctDNA detected (red, dotted) vs. not detected (blue). Median FFR time of 990 days vs. not reached (Log-Rank Test p-value 0.001); HR for ctDNA detected: 5.20, p-value 0.002). Median OS time not reached for both (Log-Rank Test p-value 0.036); HR for ctDNA detected: 2.25, p-value 0.042. E) Comparison of FFR time for patients who underwent surgery, with ctDNA detected (red, dotted) vs. not detected (blue). Median FFR time not reached for both (Log-Rank Test p-value 0.004); HR for ctDNA detected: 11.02, p-value 0.021). F) Comparison of FFR time for patients with adenocarcinoma, with ctDNA detected (red, dotted) vs. not detected (blue). Median FFR time of 686 days vs. not reached (Log-Rank Test p-value 0.0003); HR for ctDNA detected: 9.34, p-value 0.003). See fig. S4 for additional OS and CFS comparisons.

The type of treatment was strongly associated with time to event, with Log-rank p-values <0.01 for all three metrics, and significant hazard ratio (HR) values of 4.35, 2.71 and 2.01 for FFR, OS and CFS respectively with higher risk of events for patients who did not undergo surgery (Fig. 6A and B and fig. S4A). Detection of ctDNA prior to treatment was also associated with time to event, with Log-rank p-values <0.05 for all three metrics, and similar significant higher risk for patients with ctDNA detected with HRs of 5.20, 2.25 and 2.34 (Fig. 6C and D and fig. S4B). Within the largest patient subgroup (n=70), who underwent surgery, detection of ctDNA prior to treatment was associated with higher risk of events, with a significant HR of 11.02 for FFR and 2.43 for CFS, and a similar trend for OS (Fig. 6E and fig. S4C and E). In patients with adenocarcinoma, the largest histological subgroup (n=58), detection of ctDNA prior to treatment was also associated with higher risk of events, with a significant HR of 9.34, 2.89 and 2.86 for FFR, CFS and OS (Fig. 6F and fig. S4D and F).

## Discussion

In this study we analysed plasma cfDNA samples collected prior to treatment, from a cohort of 100 patients with stage I – IIIB NSCLC, using both patient-specific (INVAR) and targeted lung cancer specific mutation assays (InVisionFirst^®^-Lung) methods. The primary aim of this study was to characterise ctDNA levels in a population where detection has proven difficult in the past *(3–7)*. Through our characterisation of ctDNA levels, we now have a better understanding of the sensitivity requirements for future tools to be applied for ctDNA analysis and cancer detection in early-stage NSCLC.

The INVAR pipeline, utilising large lists of patient-specific mutations combined with signal weighting and integration, detected ctDNA in 66.7% of tested patients (60/90), including in 52.7% (29 of 55) of patients with stage I disease and in >88% of patients with stage II and III disease (16/18 and 15/17). Part of this cohort was analysed using the targeted InVisionFirst^®^-Lung assay. Detection levels of ctDNA by stage were lower using this standardised lung cancer gene panel, which is unsurprising given the evidence from other studies *(5–7, 25, 27)*. Interestingly, when combining the results from both platforms, one or both of the two platforms detected ctDNA in 94.1% (16 out of 17) of patients, an improvement over either platform alone (88.2% and 64.7% for INVAR and InVisionFirst^®^-Lung only). Our findings echo those of previous studies highlighting that the use of either a combination of different tumour markers or patient-specific approaches *(4, 7)* can improve the overall detection compared to targeted or untargeted mutation assay approaches *(28–30)*. One such example is the CancerSEEK assay which combines data from analysis of both ctDNA and protein biomarkers. Using the ctDNA component of the assay on its own, signal was detected in 22% of cases. This increased to 59% (more than 2.5-fold increase) when the protein markers were also considered *(4)*. This was particularly apparent in patients with stage I disease, where detection rates increased 10-fold from 4.3% to 43% when analysing additional markers *(4)*.

Using the distribution of ctDNA fractions across the patients in this cohort, it was possible to assess the impact on detection rates of different ctDNA detection thresholds. Setting the detection threshold at 10^−4^ had a minimal effect on ctDNA detection in patients with stage II and III disease. However, in patients with stage I disease it reduced the detection of ctDNA to less than half of those originally detected (fig. S2C). This suggests that for the baseline samples of stage I NSCLC patients, an assay would need to be able to detect ctDNA to approximately 12 ppm in order to achieve a positive call in half of the patients (Fig. 3D and Fig. 5A and fig. S2). This work echoes previous studies, showing that, especially in the context of samples with low levels of ctDNA (eg. low tumour burden or early stage of disease), the use of patient-specific mutation assays can improve analytical sensitivity and the overall rates of ctDNA detection *(5–7, 25, 27)*.

Similar to previous findings we confirmed a difference in detection rates by NSCLC subtype, showing a higher detection rate of ctDNA in patients with squamous cell carcinoma (81%) compared to patients with adenocarcinoma (59%) *(5, 7)*. This is thought to be related to differences in the biology of the two tumour types. Also similar to other work, we demonstrated a significant correlation between ctDNA levels and tumour volume *(5, 7, 24)*, and showed that this correlation is preserved at lower tumour fractions, down to a tumour volume of 0.23 cm^3^. Before this study, the smallest reported tumour volume with measurable ctDNA in lung cancer was 1.02 cm^3^ *(7)* (fig. S3A). Estimating the ctDNA fractions as a linear function of the tumour volume, for a patient with an untreated tumour with volume of 33.5 cm^3^ (equivalent to a sphere with diameter 4 cm) we estimate an average ctDNA fraction of approx. 0.08%, higher for squamous cancer (∼0.1%) and lower for adenocarcinoma (∼0.02%).

We found that detection of ctDNA prior to treatment was strongly associated with poorer patient outcome. This effect persisted in the different patient subsets. In particular, the time that patients remained free from relapse of their original primary cancer (FFR) was strongly associated with detection of ctDNA originating from that tumour. Cancer-free survival (CFS) was also significantly associated with ctDNA detection. However, the association of ctDNA detection with overall survival (OS) did not reach statistical significance (p <0.05) for the subset of patients who underwent surgery. This indicates that, while ctDNA detection prior to treatment may be a strong predictor of relapse of the primary cancer, other cancer or mortality events may occur in this at-risk population and contribute to the observed overall survival. Specifically, patients surviving lung cancer have been shown to harbour a higher risk of developing second primary cancers *(31)*.

The main challenge faced when employing personalised assays such as INVAR is the requirement of *a priori* knowledge of patient specific mutations. In our case we obtained these mutations by sequencing the exomes of matched tumour and buffy coat material obtained during surgery or from a biopsy. While approaches such as this would not be possible in a prospective study setting, we believe that our findings will support future analyses by providing a detailed characterisation of the ctDNA levels in early-stage NSCLC.

Another limitation in the use of assays requiring patient specific mutations is that they can be costly and time consuming. In order to offset the high up-front cost of INVAR, one would ideally employ patient specific assays for analysis of serially collected samples for the detection of residual disease and relapse. In this application the analysis of multiple samples for each patient with high analytical sensitivity may compensate for higher up-front cost.

Finally, despite the use of extremely sensitive assays, the data generated by this and previous studies *(5, 7)* still saw several cases with undetected ctDNA. A powerful features of INVAR is that it allows for the estimation of the expected detection threshold individually for each sample based on the number of IR obtained (Fig. 3B). With this knowledge, one could enhance sensitivity for samples of interest by identifying and targeting a larger number of patient-specific mutations (eg. through whole genome sequencing of matched tumour tissue DNA) or analyse more molecules with the given mutation list.

So far, the majority of research has focussed on patients that have already been diagnosed with cancer but ctDNA can also be a powerful tool to apply earlier at the diagnosis of cancer. Already studies are implemented that aim to diagnose cancer in the general population or in high-risk groups through ctDNA assays. Several multi-cancer early detection tests are being investigated through prospective trials *(2, 15, 32)*. If proven successful, such trials could lay the groundwork for the future application of liquid biopsies and analysis of ctDNA for the diagnosis of cancer. It will not only allow population wide screening of widely asymptomatic patients but will, more importantly, allow patients with suspected malignancies to be diagnosed sooner and could provide them with improved or earlier access to treatments appropriate for their disease. With this work we have contributed to an improvement in the general understanding of ctDNA characteristics and levels in early-stage NSCLC. This will support the development of ctDNA-based assays that will have an appropriate sensitivity to detect early-stage NSCLC.

## Data Availability

Data is available at the following EGA accession number: EGAS00001005246.

https://ega-archive.org/studies

## Acknowledgements

We would like to thank the patients and their families. We also thank the Cambridge Cancer Trials Unit - Cancer Theme, Addenbrooke’s Hospital as well as the research staff at both, Papworth and Addenbrooke’s Hospital.

## Author contributions

K. Heider, J.C.M.W. and N.R. wrote the manuscript. K. Heider generated genomic data. K. Heider, J.C.M.W., J.M., W.Q., J.W. and N.R. analysed the data and performed statistical analysis. K. Heider, J.C.M.W., F.M., C.G.S., C.M. and N.R. developed the INVAR pipeline. N.R.Q. performed imaging analysis. D.M.R. performed pathological analysis. J.C., V.R., H.K., D. Gilligan, S.V.H., R.C.R. and N.R. coordinated the clinical study and collected clinical data. D. Gale, A.R.-V., T.E., S.V.H., R.C.R. and N.R. designed the LUCID clinical study. K. Howarth and E.G. coordinated InVisionFirst^®^-Lung data and analysis. K. Heider, D. Gale and W.N.C. managed sample storage and processing. All authors reviewed and approved the manuscript.

## Data access

Sequencing data for this study is archived at the European Genome-phenome archive (EGA; www.ebi.ac.uk/ega/). To obtain access, please contact the senior author via email and complete a data access agreement with the University of Cambridge. Data is available at the following EGA accession number: EGAS00001005246.

## Methods

### Patient cohort

Samples were collected from patients enrolled to the LUCID study (REC 14/WM/1072), a prospective and observational study that enrolled patients with stage I – IIIB non-small cell lung cancer (NSCLC) undergoing treatment with curative intent, either surgery (n=70) or radio- ± chemotherapy (n=30). The primary endpoint of the study was to investigate the detection rates and levels of ctDNA at baseline in early-stage NSCLC. Informed consent was collected by a research nurse or clinician. The study was coordinated by the Cambridge Cancer Trials Unit-Cancer Theme. Additional information including patient demographics and clinical outcomes were also collected. Patient stage was classified using the 7^th^ TNM classification system, in use at the time of sample collection. Analysis was based on disease stage at diagnosis, which was available for all 100 patients. Pathological staging data was available for 70 of the 100 patients.

### Sample collection and processing

Tissue samples were obtained either from surgical specimens or diagnostic biopsies and processed as formalin-fixed paraffin-embedded (FFPE) tissue. The tissue was sectioned in 8 µm sections with one slide set aside for hematoxylin and eosin (H&E) staining to guide tumour extraction. Plasma samples were collected before treatment initiation (at baseline). For all plasma time-points, peripheral blood was collected in S-Monovette 9 mL EDTA tubes (Sarstedt). Within an hour of blood draw, samples were centrifuged at 1,600 x g for 10 minutes before undergoing another centrifugation at 20,000 x g for 10 minutes. Plasma was then stored at -80°C. A buffy coat sample was collected following removal of plasma following centrifugation of whole blood samples taken from patients prior to treatment.

### Sample extraction

For FFPE samples, the stained H&E slide was used to identify regions of high tumour cellularity, which were then macro dissected from the other tissue slides. DNA was extracted from samples using the QIAmp FFPE Tissue Kit (Qiagen) according to manufacturer’s instructions with the following modifications: DNA was incubated at 56°C and 500rpm overnight and elution was carried out by applying 20 µL ATE to the membrane twice. FFPE repair was carried out for samples containing more than 800 ng of DNA using the NEBNext^®^ FFPE DNA Repair Mix (New England Biolabs) according to the manufacturer’s instructions. DNA was extracted from up to 1 mL of Buffy coat samples either manually or using the QIAsymphony platform (Qiagen). Samples were eluted in 70 µL AE (manual extraction) or up to 200 µL elution buffer (QIAsymphony). DNA was extracted from 2 - 4 mL of plasma using the QIAsymphony robot and the DSP Circulating DNA kit (Qiagen). Samples were extracted in batches of 24 samples, including a positive and negative control to monitor extraction efficiency. FFPE and genomic DNA were quantified using a dsDNA broad range assay on the Qubit fluorimeter (ThermoFisher Scientific). DNA in plasma samples was quantified using a digital PCR with 55 cycles on a Biomark HD (Fluidigm) using a dual-labelled probe targeting a 65bp region of the *RPP30* gene (Sigma Aldrich) *(25)*.

### Library preparation

Using the Covaris LE220 (Covaris) according to manufacturer’s instructions, tumour and buffy coat DNA were sheared to a fragment length of 200 bp. 15 µL volumes and the 8 microTUBE-15 AFA Beads Strip V2 were used and fragmentation patterns of random samples were checked using a Bioanalyser (Agilent). A total of 100 ng (tumour) and 50 ng (buffy coat) of sheared DNA were used for library preparation with the ThruPLEX DNA-seq kit (Rubicon). The number of library cycles was adjusted to the sample input according to the manufacturer’s recommendations.

Up to 15 ng of plasma DNA were used for library preparation with the ThruPLEX Tag-seq (Rubicon) or SureSelect XTHS kit (Agilent). Depending on input amount, the number of amplification cycles was varied according to the recommendations from the manufacturer. After library preparation, qPCR (NEBNext^®^ Library Quant Kit for Illumina^®^ in the ROX low setting, New England Biolabs) and Bioanalyser or TapeStation (both Agilent) were used to determine library concentration and size.

### Exome capture of tumour and buffy coat samples

The Illumina TruSeq kit with a 45 Mbp bait set (Illumina) was used for the exome capture of tumour and buffy coat samples after library preparation. Keeping tumour and buffy coat DNA separate, 250 ng of each library were multiplexed in three-plex reactions. To ensure compatibility with the ThruPLEX libraries, 1 µL of i5 and i7 TruSeq HT xGen universal blocking oligos (IDT) were added at the hybridisation step and the volume of CT3 buffer was adjusted to 51 µL. Samples underwent two rounds of hybridisation, each lasting for 24 hours. After exome capture, sample QC was performed as described above and sequencing was carried out using a HiSeq4000 (Illumina).

### Mutation calling in tumour tissue

Tumour mutation calling was carried out on two different batches of patient samples. For the first batch of FFPE tumour biopsies, mutation calling was performed with Mutect2 with the default settings: --cosmic v77/cosmic.vcf and --dbsnp v147/dbsnp.vcf. To maximise the number of mutations retained, all variants achieving Mutect2 pass were retained. Mutation calls were then filtered as follows:

1. Buffy coat mutant allele fraction equals zero
2. Mutation not in homologous region
3. Mutation not at a multiallelic locus
4. 1000 Genomes ALL and EUR frequency equals zero
5. A minimum unique tumour depth of 5

For the second batch of patient samples, mutations were identified using Mutect2, VarDict and Freebayes. The same filters as described above were applied to this cohort. In addition to retaining mutations passing Mutect2, mutations identified by both Freebayes and VarDict were also retained, thereby increasing our total number of targetable mutations. In some cases, multiple tumour regions were available for the same patient so mutation calling was carried out on the individual tumour regions as well as from the merged bam files of both regions.

### Design of hybrid-custom capture panels, target capture and sequencing of plasma samples

Following mutation calling, custom capture panels were designed using SureDesign (Agilent). Mutation lists from 19 to 35 patients were combined per capture panel and a 1x tiling density and balanced boosting was used. 120 bp RNA baits were used for the panels, which varied in size from 2.138Mbp to 2.987Mbp.

Target-enriched libraries were generated by performing hybrid-capture of the plasma cell-free DNA libraries, in singleplex or two-plex up to a total input of 1000 ng. SureSelect XT and SureSelect XTHS hybrid capture was carried out according to the manufacturer’s instructions. For ThruPLEX Tag-Seq libraries, i5 and i7 blocking oligos (IDT) were added based on the manufacturer’s recommendations. Captured libraries underwent 13 cycles of post-capture amplification and underwent the same QC as described above. Target-enriched libraries were sequenced on the HiSeq4000 platform (Illumina).

### Read collapsing on plasma sequencing data

Known 5’ and 3’ adapter sequences were specified in a separate FASTA file and removed using Cutadapt v1.9.1. Using BWA-mem v0.7.13 with a seed length of 19, the trimmed FASTQ files were aligned to the UCSC hg19 genome. All samples underwent read collapsing using CONNOR *(33)* with a consensus threshold of 90% (-f flag of 0.9) and a minimum family size of 2 (-s flag of 2).

### Plasma analysis using the INVAR pipeline

Plasma custom-capture sequencing data was analysed using the INVAR pipeline, which was described previously *(16)*. The following minor modification was made when running the pipeline. As this was an early stage cohort with few mutant fragments expected in each sample, dasta from another early stage cohort was added to better characterise the fragment size distribution of mutant and wildtype reads. This combined size distribution file was used when weighting mutant fragments based on their fragment length.

### Plasma analysis using the InVisionFirst^®^-Lung assay

Up to 4 mL of plasma were sent to Inivata, where samples were extracted and analysed using the InVisionFirst^®^-Lung assay *(17, 18)*. The assay utilises the eTAm-Seq™ technology, which is based on the previously developed TAm-Seq method *(25)*. Sequencing libraries were prepared using a two-step PCR amplification protocol with amplicons (72bp-154bp in length) covering 10.61kb across 36 cancer related genes. Mutations detected and allele fractions for each were reported. If more than one mutation was detected in a patient, the ctDNA allele fraction was calculated as the mean allele fraction of the detected mutations. Where available, mutations detected in plasma were compared to matched buffy coat sequencing data to remove any potential germline mutations.

### Summary statistics analysis

Standard descriptive summary statistics were used for the data summaries. Continuous variables were summarised as number of patients, mean (standard deviation - SD), median (minimum, maximum) and interquartile range (IQR) whilst categorical variables were summarised as percentage. ctDNA detection rates were analysed with respect to different clinical features using Clopper-Pearson 95% confidence interval (CI).

### Volumetric tumour analysis

Volumetric analysis of stage I and II lung cancers was performed using the Siemens Syngo Via MM Oncology™ imaging software tool (Siemens, Erlangen, Germany). This allowed automated tumour segmentation of DICOM compatible CT images for three-dimensional volumetric assessment. For lesions that abutted the mediastinum, vascular structures or chest wall, a manual nudge modification of the volume of interest was performed to ensure optimal volumetric analysis.

## Supplementary figures

**Fig. S1:**
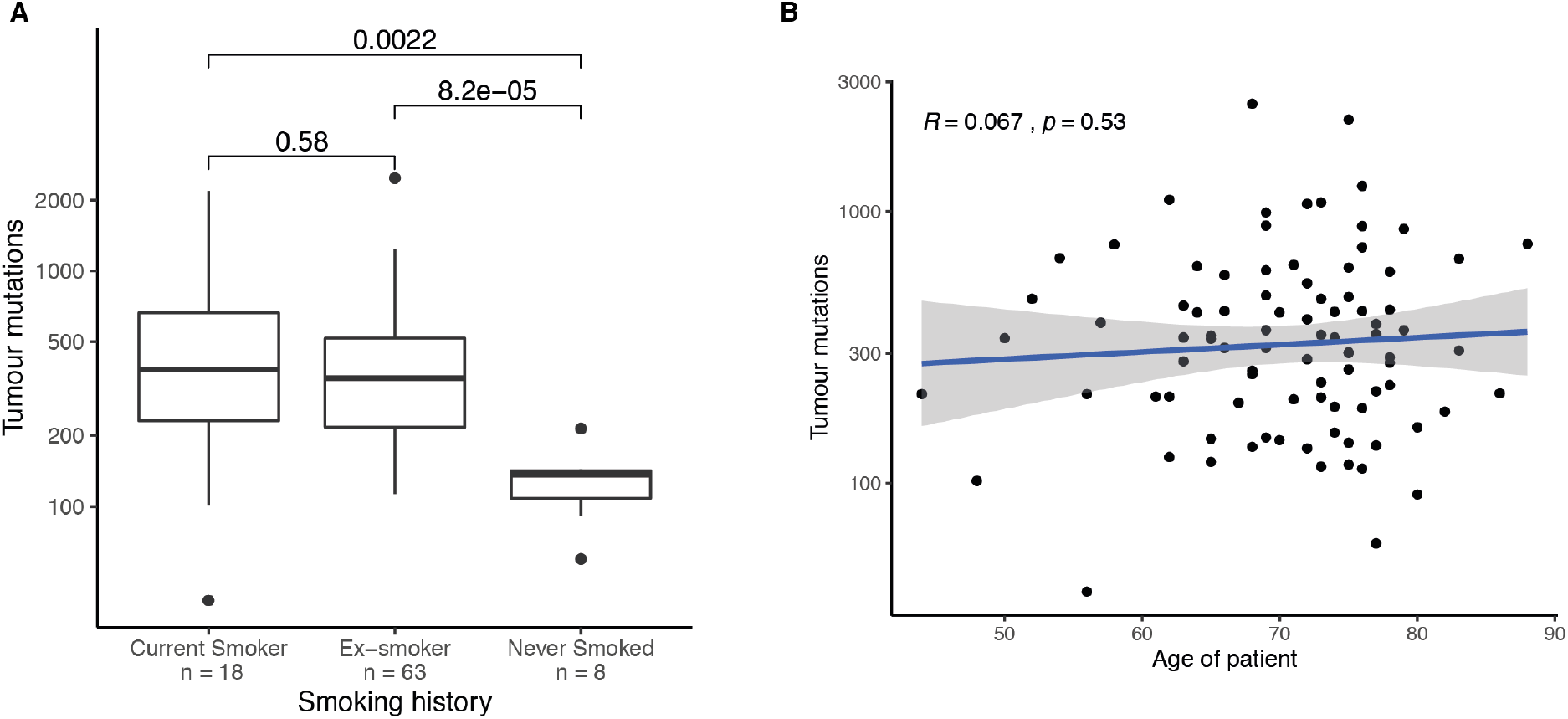
(A) Distribution of the number of mutations identified in exome-wide sequencing of tumour samples, for patients with different smoking status/history. Current and former smokers had significantly more mutations identified in their tumour tissue samples compared to never smokers. (B) Tumour mutations per patient plotted against the age of the patient. No significant correlation was observed.

**Fig. S2:**
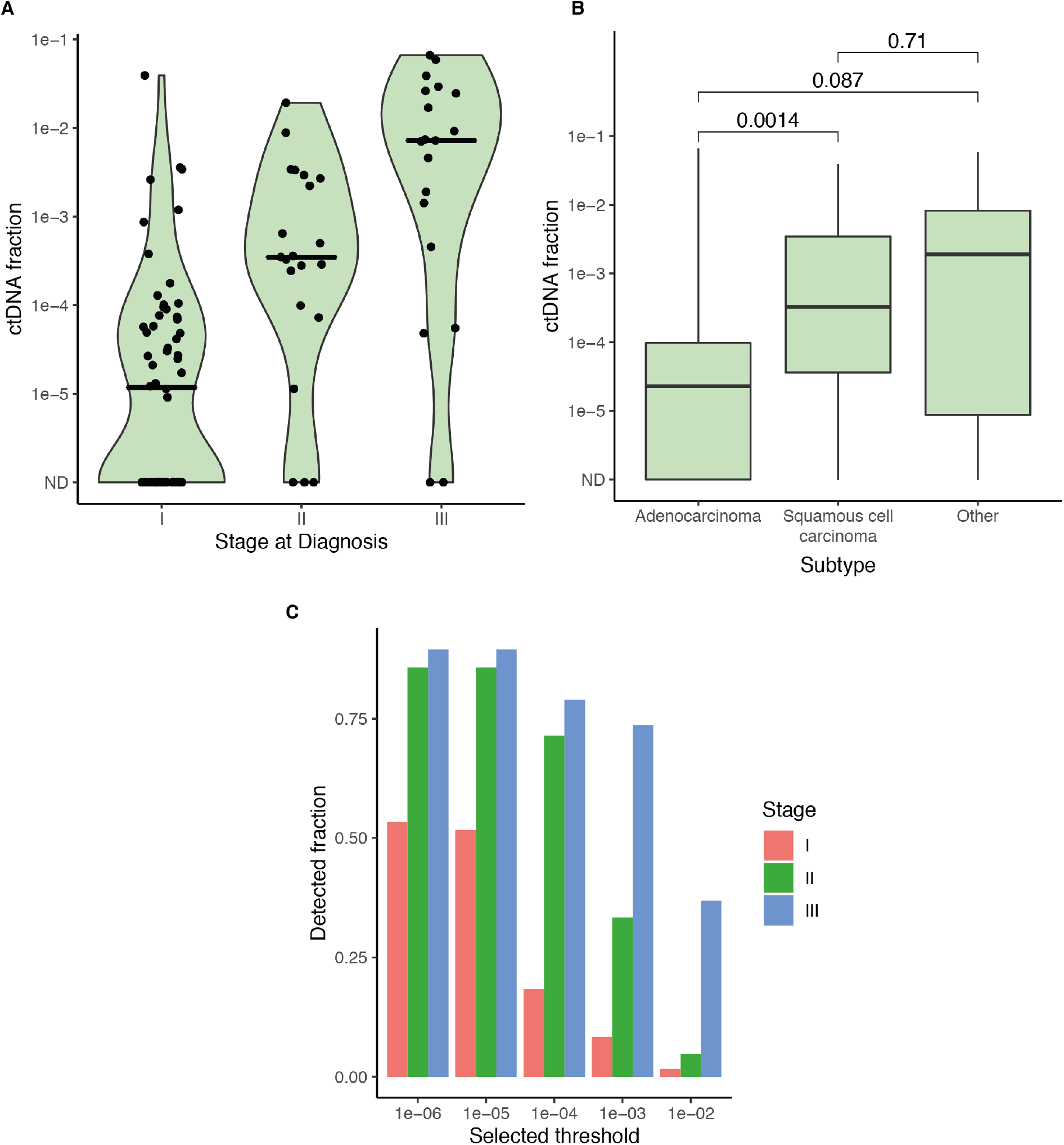
(A) Violin plot of ctDNA fraction by stage of disease at diagnosis shows increasing ctDNA fraction with increasing stage. Horizontal lines indicate median levels. Data includes all 100 patients. (B) Boxplot of ctDNA fraction and cancer subtype shows significantly lower fractions of ctDNA prior to treatment in patients with adenocarcinoma. (C) The fraction of plasma samples with ctDNA detected out of all 100 patients in the study, when different thresholds for detection are imposed ranging from ctDNA fraction of 1×10^−6^ (which includes all samples detected in this study) to fractional concentration of 1×10^−2^. Detection rates decreased with increasing thresholds; this decrease was most notable up to ctDNA fraction of 1×10^−3^ for patients with early-stage disease.

**Fig. S3:**
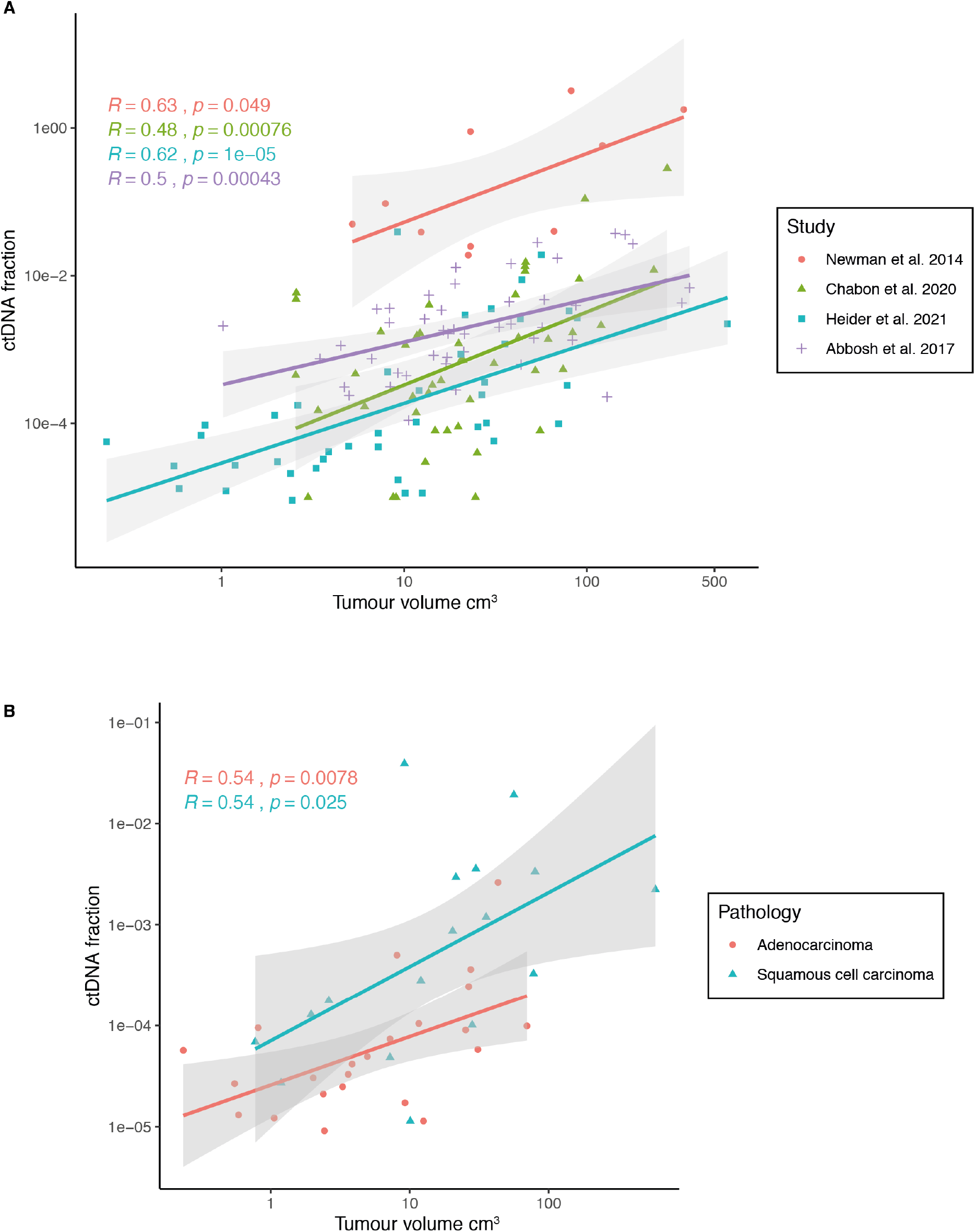
(A) Correlation between ctDNA and tumour volume in the present study and recently published reports. While all studies show a significant correlation between ctDNA and tumour volume, the present study also detected and quantified ctDNA fractions in patients with tumour volumes below 1 cm^3^. B) Comparing ctDNA fractions in patients with squamous cancers (blue triangle) and adenocarcinomas (red dots) shows that for the same tumour volume, ctDNA fractions are higher in squamous cancers vs. adenocarcinomas.

**Fig. S4:**
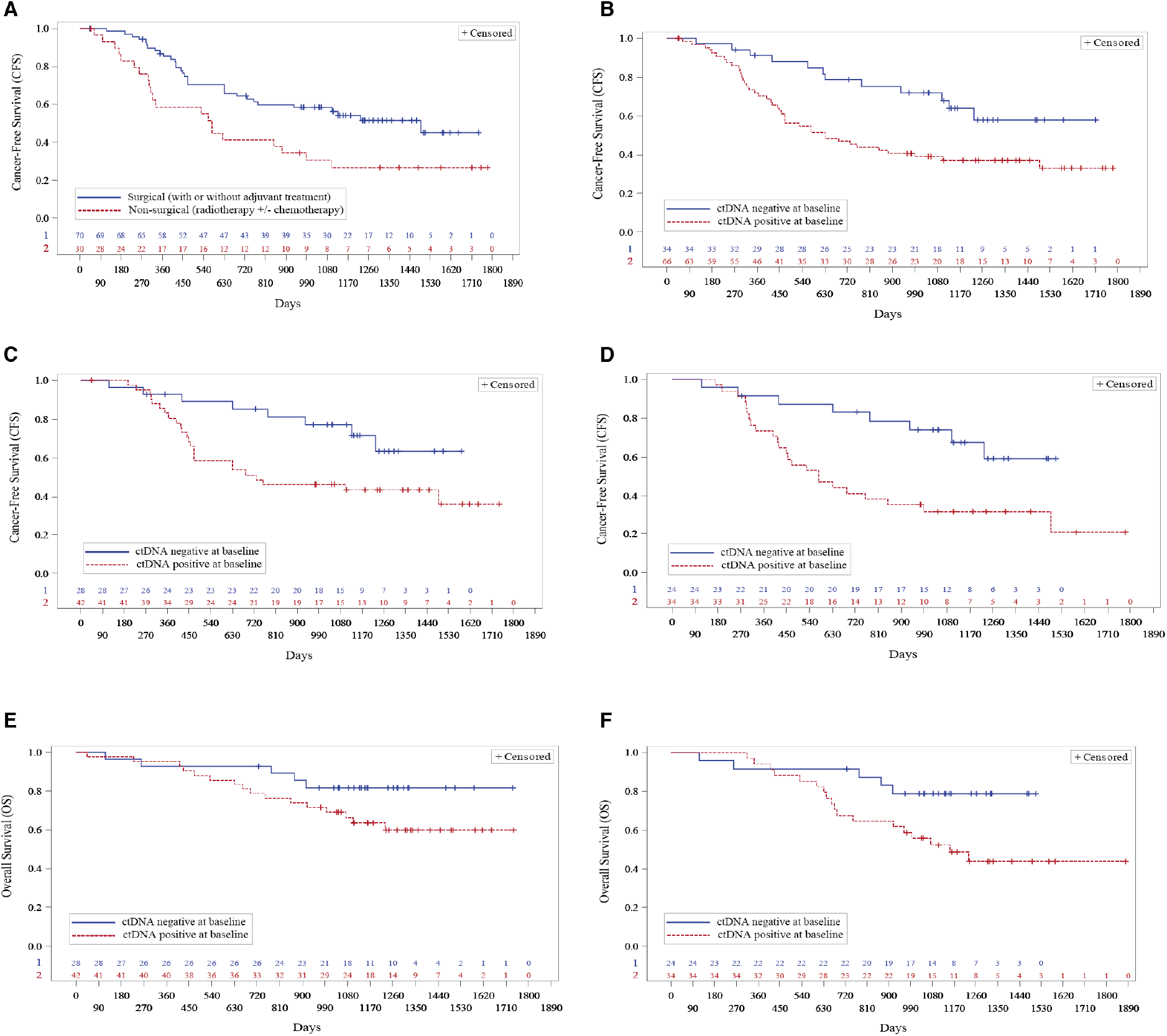
Kaplan-Meier plots showing overall survival (**OS**) and cancer-free survival considering all cancer or death events (**CFS**). (A) Comparison of CFS for patients who underwent non-surgical treatment (red, dotted) vs. surgical treatment (blue). Median CFS of 576 days vs. 1490 days (Log-Rank Test p-value 0.009); HR for non-surgical treatment: 2.05, p-value 0.011). (B) Comparison of CFS for all patients, with ctDNA detected (red, dotted) vs. not detected (blue). Median CFS of 632 days vs. not reached (Log-Rank Test p-value 0.008); HR for ctDNA detected: 2.34, p-value 0.010). (C) Comparison of CFS for patients who underwent surgery, with ctDNA detected (red, dotted) vs. not detected (blue). Median CFS 731 days vs. not reached (Log-Rank Test p-value 0.026); HR for ctDNA detected: 2.43, p-value 0.031). (D) Comparison of CFS for patients with adenocarcinoma, with ctDNA detected (red, dotted) vs. not detected (blue). Median CFS 577 days vs. not reached (Log-Rank Test p-value 0.007); HR for ctDNA detected: 2.89, p-value 0.010). (E) Comparison of OS for patients who underwent surgery, with ctDNA detected (red, dotted) vs. not detected (blue). Median OS not reached for both (Log-Rank Test p-value 0.099); HR for ctDNA detected: 2.27, p-value 0.109. (F) Comparison of OS for patients with adenocarcinoma, with ctDNA detected (red, dotted) vs. not detected (blue). Median OS 1154 days vs. not reached (Log-Rank Test p-value 0.030); HR for ctDNA detected: 2.86, p-value 0.038).

